# Co-designing a theory-informed, multi-component intervention to increase vaccine uptake with Congolese migrants: a qualitative, community-based participatory research study

**DOI:** 10.1101/2023.05.30.23290568

**Authors:** Alison F Crawshaw, Lusau Mimi Kitoko, Sarah Nkembi, Laura Muzinga Lutumba, Caroline Hickey, Anna Deal, Jessica Carter, Felicity Knights, Tushna Vandrevala, Alice S Forster, Sally Hargreaves

## Abstract

**Introduction:** Inequitable uptake of routine and COVID-19 vaccinations has been documented among intersectionally marginalised populations, including migrants, and attributed to issues of mistrust, access, and low vaccine confidence. Novel approaches which seek to share power, build trust and co-design tailored interventions with marginalised or underserved communities must be explored, to promote equitable engagement with vaccination and other health interventions.

**Methods:** A theory-informed, qualitative, community-based participatory research study, designed and led by a community-academic partnership, which aimed to understand decision-making related to COVID-19 vaccination among Congolese migrants in the UK and co-design a tailored intervention to strengthen their vaccine uptake (2021-2022). Barriers and facilitators to COVID-19 vaccination, information and communication preferences, and intervention suggestions were explored through qualitative in-depth interviews with Congolese migrants, thematically analysed, and mapped to the theoretical domains framework (TDF) and the Capability, Opportunity, Motivation, Behaviour (COM-B) model to identify target behaviours and strategies to include in interventions. Workshops were done in partnership with Congolese migrants to co-design and tailor interventions.

**Results:** 32 Congolese adult migrants (foreign-born and living in UK; 24 (75%) women, mean 14.3 [SD 7.5] years in UK, mean age 52.6 [SD 11.0] years) took part in in-depth interviews and 16 (same sample) took part in co-design workshops. We identified 14 barriers and 10 facilitators to COVID-19 vaccination; most barrier data related to four TDF domains (beliefs about consequences; emotion; social influences; environmental context and resources), and the behavioural diagnosis concluded interventions should target improving psychological capability, reflective and automatic motivations, and social opportunities. Strategies included behaviour change techniques based on education, persuasion, modelling, enablement, and environmental restructuring, which resulted in a co-designed intervention comprising community-led workshops, COVID-19 vaccination plays and posters. Findings and interventions were disseminated through a community celebration event.

**Conclusions:** Our study demonstrates how behavioural theory can be applied to co-designing tailored interventions with marginalised migrant communities through a participatory research paradigm to address a range of health issues and inequalities. Future research should build on this empowering approach, with the goal of developing more sensitive vaccination services and interventions which respond to migrant communities’ unique cultural needs and realities.

**Patient or public contribution:** Patient and public involvement (PPI) were embedded in the participatory study design and approach. An independent PPI board comprising five adult migrants with lived experience of accessing healthcare in the UK were also consulted at significant points over the course of the study.

**Practitioner points:**

- Research has shown that migrants experience a range of health and vaccination inequalities but are not well included in health research nor the design of interventions to address these. Using community-based participatory methods, we demonstrated that underserved communities, such as migrants, are resilient, resourceful, and use community assets to find real-world solutions to their health needs.
- Our approach shows how practitioners can adapt and use behavioural theory and design thinking within a participatory research paradigm to meaningfully involve underserved populations in co-designing acceptable and culturally relevant health interventions to address a range of health issues and inequalities.

## INTRODUCTION

Vaccination is one of the world’s most cost effective and successful public health interventions and is essential to reducing deaths and improving health outcomes caused by serious infectious diseases. Faced with the COVID-19 pandemic, scientists and governments rapidly set about developing and distributing safe and effective vaccines for COVID-19 to help bring the pandemic under control and protect populations. However, the success of vaccine-based protection measures hinges on high population uptake and coverage. Monitoring of the COVID-19 vaccination roll-out in high income countries revealed stark gaps and discrepancies in COVID-19 vaccine uptake particularly affecting intersectionally marginalised populations, including migrants (1–8). In the UK and Europe, several studies have suggested migrants are also an under-immunised group for routine vaccinations, with few systems in place to engage and catch-up older age groups (9–12). Barriers include poor access to vaccines despite availability, low confidence in vaccine safety and effectiveness, and low trust in public institutions and the wider health system (6, 10, 13, 14). Many of these same populations also suffered disproportionately worse health and economic outcomes because of the pandemic (15, 16).

Health inequalities can be linked to wider social inequalities, including broader environmental, social, and economic factors. Globally, COVID-19 exacerbated inequalities experienced by some migrants and ethnically minoritised groups and highlighted the structural violence embedded within society (17, 18). Along with hostile immigration policies, institutional racism, and xenophobia, the medical establishment has a long history of exploiting and mistreating black and some ethnically minoritised populations (19, 20). This is reflected in poorer health outcomes for and between many of these groups compared to white groups. For example, rates of infant and maternal mortality, cardiovascular disease and diabetes are higher among Black and South Asian groups. The effects of this legacy and wider context on trust were also evident in widely reported conspiracy theories about population control and concerns of being used as ‘guinea pigs’ in the COVID-19 vaccination drive, posing major barriers to vaccine uptake (1, 21, 22). Muddled and inconsistent messaging and a lack of leadership from Heads of State during acute phases of the pandemic also likely contributed to lower trust in the health system and allowed misinformation to thrive (23), particularly among migrant and ethnically minoritised groups. There were also clear information barriers for those with limited English language proficiency and failure of governments to adequately adapt and disseminate essential messaging to diverse populations (24). Although governments later took steps to physically widen access to COVID-19 vaccination for excluded groups (25, 26), these actions were not enough to repair their already eroded trust in public institutions and authorities. As we now begin to move from pandemic to more endemic stages of COVID-19, it is essential that we do not lose sight of the inequities highlighted nor the momentum needed to tackle them. This is important not only to improve COVID-19 vaccine equity, but to improve the reach of routine vaccination programmes and improve health outcomes for affected populations more broadly. The King’s Fund recently stated that “a cross-government strategy for reducing health inequalities and addressing the diverse health needs of all groups at risk of poor health and high mortality has never been more urgent” (27). This must be done sensitively, in a way which considers pre-existing structures of oppression and mistrust and adequately accounts for populations’ unique realities, lived experiences and diversity.

Various approaches based on behavioural insights theory have been used to increase uptake of routine and other more established vaccinations, including the World Health Organization’s (WHO) Tailoring Immunisations Programme (TIP). WHO TIP recognises that individual behavioural, contextual, social, and societal factors influence vaccination uptake and provide a framework to understand and address these (28). The intervention development phase of TIP is based on the well-known Capability, Opportunity, Motivation, Behaviour (COM-B) model of behaviour change and its corresponding theoretical domains framework (TDF), which have been combined to form the Behaviour Change Wheel (BCW) (29–31). The TDF is an integrated theoretical framework comprising 14 validated domains (see Figure 1) that can be used to identify the determinants of a given behaviour (e.g. vaccination). The COM-B model assigns these domains to three interacting factors which predict behaviour: capability, opportunity, and motivation. These sources of behaviour can then be used with the next layer of the BCW to identify potentially relevant intervention functions. Strengths of the WHO TIP approach are that it helps to build in-depth mutual understanding and trust between a variety of stakeholders, recognises the complex mechanisms that influence vaccination behaviour, and goes beyond diagnosis of barriers to implementation of interventions supporting change. A limitation is that it typically takes place within a traditional research paradigm, where studies are designed and implemented by academics and research is done “on” rather than “with” communities. This power dynamic can be harmful and oppressive to communities, particularly those who are already minoritised or marginalised, and perpetuate inequities. It can also lead to inauthentic participation and under-representation of these groups in research (32).

**Figure 1.**
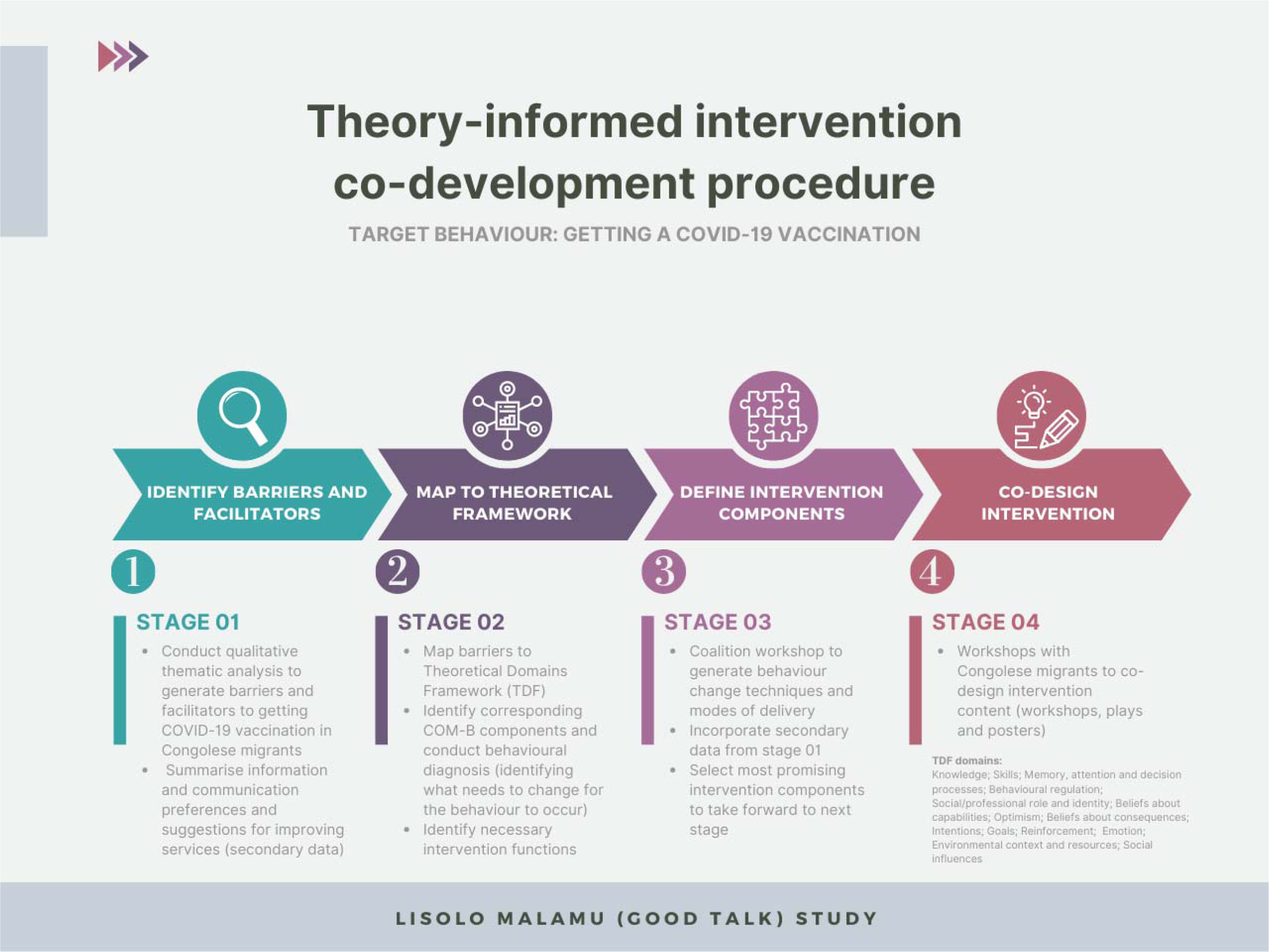
The four stages of the theory-informed intervention co-development procedure: identify barriers and facilitators; map to theoretical framework; define intervention components; co-design interventions. Target behaviour was getting a COVID-19 vaccination.

In contrast, a participatory research paradigm directly considers power asymmetries and histories of oppression, gives value to the subjectivity of lived experience, and actively involves individuals affected by the issue being studied as equal partners in the research process. Participatory research leads to knowledge that is locally situated and context-specific, which is important for generating workable solutions to existing problems (33). In addition to enhancing community empowerment, it is argued that engaging communities in this way can advance the rigor, relevance and reach of research (34). To date, there have been shortcomings in the meaningful involvement of migrants in health research (35), which we see as an opportunity for improvement. The resurgence of interest in participatory research provides an exciting opportunity to rethink existing approaches to addressing vaccine inequities and involving migrant populations in research through an inclusive, collaborative, and community-centred lens, and may advance efforts to close the global immunisation gap.

Inspired by the existing approaches to addressing this challenge and their shortcomings, we recognised that a novel contribution at the time of this study’s inception (early 2021) would be a study that could:

- Conduct careful in-depth research with a specific underserved migrant population, considering the complex mechanisms influencing their vaccination attitudes, beliefs and behaviours;
- Translate these findings into a theory-informed tailored intervention;
- Share power, build trust, and meaningfully involve members of the target population in all stages of the study and intervention co-design through a collaborative, participatory research approach.

We therefore constructed this study with Congolese migrants in the UK to explore the individual behavioural, social, societal, and contextual factors influencing their attitudes, beliefs and behaviours related to COVID-19 vaccination, and used behavioural theory, design thinking and a community-based participatory research approach (CBPR) to co-design an intervention to strengthen their COVID-19 vaccine uptake, which was evaluated. Although specific to COVID-19 vaccination and Congolese migrants, we believe the approach used (36) is relevant to engaging with other underserved populations and developing tailored interventions to increase uptake of other routine, seasonal, and catch-up vaccinations, as well as other health interventions.

## METHODS

### Study design and setting

This qualitative study was conducted as part of a CBPR study investigating routine, catch-up and COVID-19 vaccination that was designed and led by a community-academic coalition and conducted in partnership with Congolese migrants in a diverse borough of London, UK. Methods are described in detail in a published protocol (36), including context for this study, details about forming the coalition, study population, data collection and analysis. Additional methods used and variations in procedures are described below. All study resources and expenses were paid for by grants awarded to the [institution name redacted] research team. Participants were financially compensated for participation using vouchers and reimbursed in cash for travel costs. Non-academic coalition members were paid for their time based on NIHR INVOLVE guidance (37) and [organisation name redacted] and [organisation name redacted] received financial donations to support their running, in additional to non-financial contributions (e.g. skills-based training) (36).

The aim of this study was to co-design a tailored intervention to strengthen COVID-19 vaccine uptake with Congolese migrants living in London. Migrants were defined as non-UK born. We were interested to understand the decision-making related to COVID-19 vaccine uptake and specified our target behaviour (as recommended by Michie and colleagues (29)) as ‘getting a COVID-19 vaccination’, though this behaviour was not a requirement. The four stages of intervention development are described below and shown in Figure 1. We used qualitative and co-design methods underpinned by the Theoretical Domains Framework (TDF), COM-B model and Behaviour Change Wheel (BCW) as our theoretical framework. These three interlinking frameworks provide a means to theoretically and systematically understand and change behaviour (38). We chose them because they are comprehensive, theory-informed, and have been independently validated and used successfully in health and implementation research (39, 40).

Qualitative in-depth interviews were conducted by four members of the study coalition (1 academic researcher and 3 Congolese women trained as peer researchers) at a local community centre across 3 days in January – March 2022. Interviews were conducted in Lingala, French or English depending on the participant’s preference. Interviews were audio recorded, transcribed verbatim and pseudonymised, and translated by a professional translator, where required. Field notes were manually added to transcripts and data were analysed by hand and using NVivo software. Two co-design workshops (with different participants) were conducted by five members of the coalition [initials redacted] at a local community centre in May 2022 in Lingala and English. Optional, open-ended evaluation forms were collected after the interviews and workshops and the text qualitatively analysed using a simple framework matrix. A community celebration and presentation of key findings was held in July 2022.

### Sampling and recruitment of participants for in-depth interviews and co-design workshops

We aimed to recruit around 30 participants for the in-depth interviews and 6-8 workshop participants, who were born in the Democratic Republic of Congo (DRC), residing in the UK, and aged over 18 years (see inclusion criteria in Table 1 and in our published protocol for further detail (36)).

**Table 1.** Inclusion and exclusion criteria of study participants.

| Inclusion criteria | Exclusion criteria |
| --- | --- |
| Born in the Democratic Republic of Congo (DRC).<br>Aged 18 or above.<br>Currently residing in the UK.<br>Willing and able to give informed consent. | Not migrant as per earlier definition.<br>Not born in the DRC.<br>Below the age of 18.<br>Temporarily in the UK for holiday, visiting friends/relatives, or other reasons.<br>Lacking capacity to consent, as determined by the mental capacity act framework. |

Participants were recruited by the three Congolese members [initials redacted] of the study coalition, by word-of-mouth, flyers co-designed by the coalition, and snowball sampling techniques. All participants received a participant information sheet (PIS) explaining the study and their rights, which was also explained to them verbally, and had the opportunity to ask questions and decide whether to participate. The PIS explained that the study involved 2 parts, an in-depth interview lasting around 30-45 minutes and a co-design workshop lasting around 2 hours, and that they could take part in neither, both or just one part. During the in-depth interview consent process, participants could indicate whether they would like to be invited to join a workshop. Those who indicated ‘yes’ were followed up after the in-depth interviews by one of the three Congolese coalition members, who explained the workshop and invited them to attend. Written informed consent was obtained prior to starting each workshop.

### Ethical approval and consent to participate

Ethics was granted by the St George’s University of London Research Ethics Committee (REC reference 2021.0128). All participants provided informed consent and were older than 18 years at the time of recruitment to the study.

### Breakdown of study costs

In total, this study cost approximately £17,500 to conduct, not including academic staff time. This included £7000 on general project spend (coalition member payments and expenses, participant vouchers and expenses, venue hire, catering and entertainment for end-of-study celebration event, stationery and other materials, professional artist hire), £4500 in one-off donations to non-academic partners, and £6000 on translation and transcription costs. Translation costs were an unforeseen expense due to Lingala being considered a rare language by translation companies and priced accordingly. Ultimately, an independent professional translator from the London Congolese community was used, who offered a more competitive rate than established translation companies.

### Intervention development procedure

The target behaviour specified was ‘getting a COVID-19 vaccination’. Intervention development involved 4 stages: thematic analysis (41) and coding of qualitative interview data; mapping of barriers and facilitators to the TDF, COM-B and BCW intervention functions; coalition workshop to ideate behaviour change techniques and select potential intervention candidates to carry forward to co-design workshops; and co-design workshops with Congolese participants to co-design intervention content set out by the previous stage (Figure 1).

### Stage 1-thematic analysis and coding of qualitative interview data

The full qualitative dataset explored perceptions and experiences of routine, catch-up and COVID-19 vaccination and was analysed using a reflexive thematic analysis (42) which will be reported elsewhere (manuscript *in preparation*). This analysis focused on data regarding COVID-19 vaccination and health-related beliefs, attitudes, and behaviours (see Figure 1). It was a systematic process, with initial data familiarisation and analysis done collaboratively: [initials redacted] repeatedly read the transcripts and critically engaged with the data, writing personal notes and questions to discuss with the coalition who had also immersed themselves in the data (familiarisation). For pragmatic reasons, data were first coded deductively, using broad pre-defined codes (barriers, facilitators, information and communication preferences, suggestions for interventions). New codes were then generated inductively, iterated on, and refined progressively through multiple rounds of coding and collaborative analysis sessions involving the coalition. Codes were then organised into topic summaries to aid initial theme generation, including barriers and facilitators to COVID-19 vaccination and information and communication preferences, which included preferred formats, trusted sources, information channels and meeting points, and messaging contents.

### Stage 2 – mapping of barriers and facilitators to the TDF, COM-B and BCW intervention functions

Barrier concepts were mapped to the 14-domain TDF framework (30, 43), (TDF domains are listed in Figure 1). Mapping was an iterative and subjective process that involved discussions between the coalition [initials redacted] until consensus was reached. Michie et al. 2014 have developed numerous resources to aid intervention development that link the TDF domains to COM-B components and BCW intervention functions (31). Using their expert consensus matrix (31) and resources, we identified the corresponding COM-B components, conducted a behavioural diagnosis (a process to identify changes needed for the target behaviour of COVID-19 vaccination to occur) and identified potential intervention functions likely to be effective in bringing about those changes.

### Stage 3 – coalition workshop to generate behaviour change techniques, modes of delivery and select interventions

Working from the behavioural diagnosis, we [initials redacted] selected 5 intervention functions to focus on for the intervention development for practical reasons; these were based on the functions’ recurrence across the framework (representing their applicability to key concepts) and suitability based on APEASE criteria (31). We then developed a series of ‘How Might We…?’ (HMW) questions from the behavioural diagnosis, borrowing a method traditionally used in design thinking to reframe a problem and generate creative solutions (44), and which has not been applied in this way before (to the best of our knowledge). A matrix linking the HMW questions and their corresponding intervention functions was created as a prompt for the ideation sessions (Table S2). In small groups, we spent 5 minutes rapidly ideating (brainstorming) possible behaviour change techniques (BCTs), modes of delivery and content (‘intervention components’) on post-it notes to address each HMW question. Suggestions from participants obtained during IDIs were also written on post-its and all ideas were collated under each HMW question. After each ideation session, we discussed, refined, and evaluated ideas collectively, taking into consideration the information, communication and cultural preferences identified in stage 1. We then selected the three most promising intervention components (based on perceived importance of barriers, appropriateness and feasibility of the approach, and impact versus effort) to take forward for development in the co-design workshops with members of the target population. Benefits of the coalition doing this exercise are that we were all immersed in the data, and our coalition members from the target population were especially familiar with cultural preferences and values ensuring these could be adequately reflected in the ideas, discussions and chosen components.

### Stage 4 – intervention co-design workshops

Two, two-hour co-design workshops were held with Congolese migrants and led by the Congolese coalition members [initials redacted], with facilitation support from [initials redacted]. Participants who expressed interest in participating in the workshops were randomly selected and invited for up to a total of 16 participants (8 per workshop; group size chosen based on a discussion of what would be culturally acceptable, manageable, inclusive and productive). A local artist recorded visual minutes.

The workshops began with introductions and ice breakers, followed by longer activities to co-develop interventions in breakout groups. The coalition decided that both workshops should address the first intervention components (being more informational), and then each take one of the two remaining intervention components (being more creative). Coalition members facilitated the breakout groups and used tools and resources developed prior to the workshops to encourage the participants to think creatively and tailor their interventions and content. We also shared examples of real public health campaigns (e.g. for HIV, Ebola, etc) to inspire participants. The groups worked on each intervention for approximately 30 minutes, before returning to present and discuss their designs in the round. At the end of the activity, all participants discussed and agreed on the best ways to disseminate their interventions to the community and voted for the most promising design(s). A community celebration and research dissemination event was later held to mark the end of the study.

## RESULTS

Results are presented in three parts, following the stages outlined in the methods: i) barriers and facilitators to COVID-19 vaccination, information and communication preferences, and suggestions for improvement (stage 1); ii) behavioural diagnosis and selection of interventions (stages 2 & 3); and iii) outputs of the co-design workshops (stage 4).

We conducted 32 interviews with Congolese migrants and 2 co-design workshops with 16 participants (8 per workshop). Descriptive characteristics of the qualitative interview participants (n=32) are shown in Table 2 and described briefly here. Co-design participants were drawn from this sample. Most (75%) of the interview participants were female, had a mean age of 52.6 years (SD: 11 years), and had lived in the UK for an average (mean) 14.3 years (SD: 7.5 years). We expanded our inclusion criteria to include two Congolese-identifying but Angolan-born participants, recognising the limitations of our original categories. Most participants spoke Lingala (88%) or French (63%); few spoke English (31%) and 47% considered themselves to have limited English proficiency (unable to read or write). All (100%) were registered with a GP. Interviewees were asked their COVID-19 vaccination status and number of doses received at the time of their interview (conducted January – March 2022). 4 (13%) answered ‘unvaccinated/0 doses’, 18 (56%) answered ‘1-2 doses’, 10 (31%) answered ‘3 or more doses’, and 1 (3%) answered ‘uncertain’. In the co-design workshops, there was an almost even sex distribution (4 women, 4 men in workshop 1; 3 women, 5 men in workshop 2).

**Table 2:**
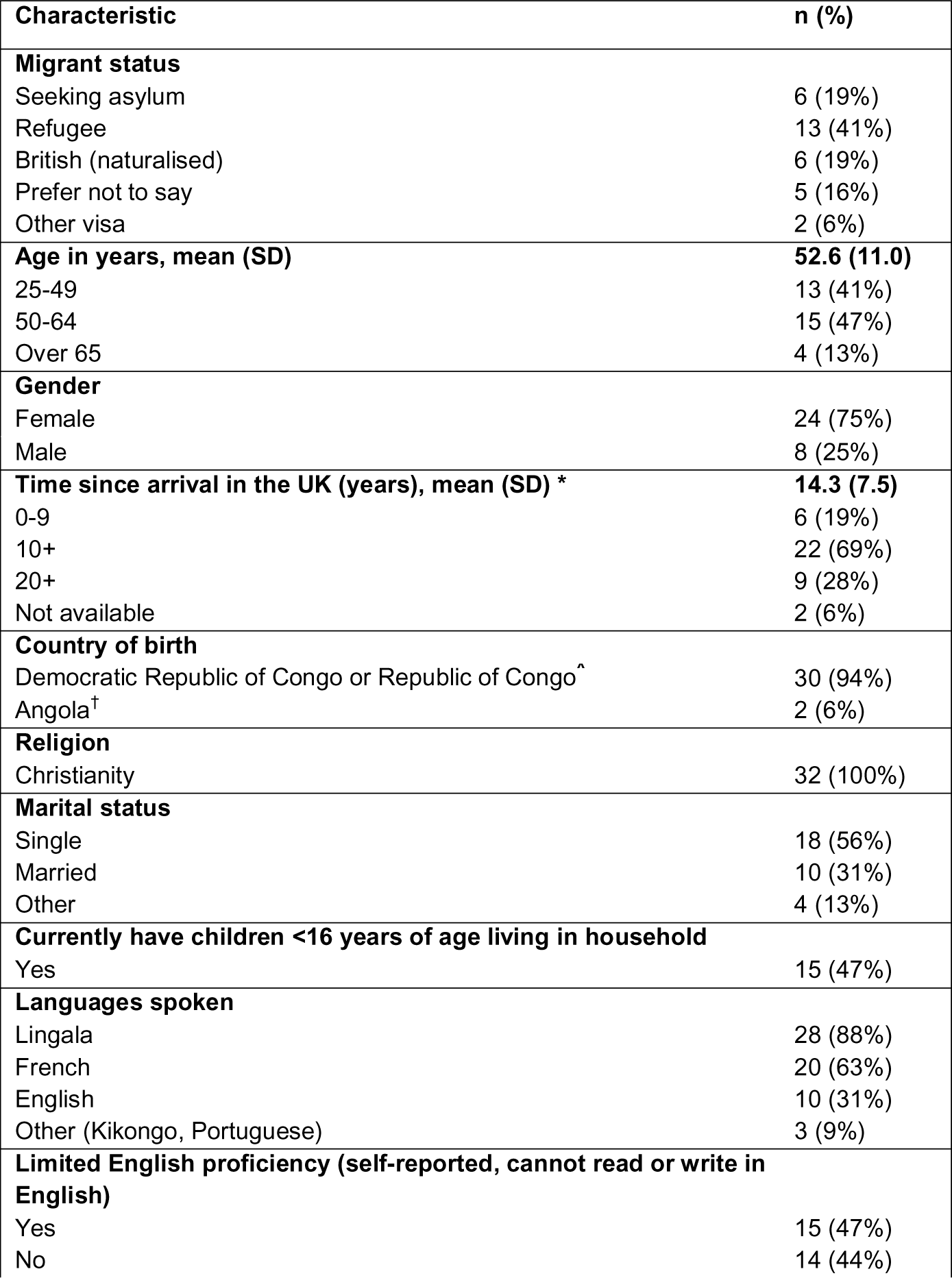
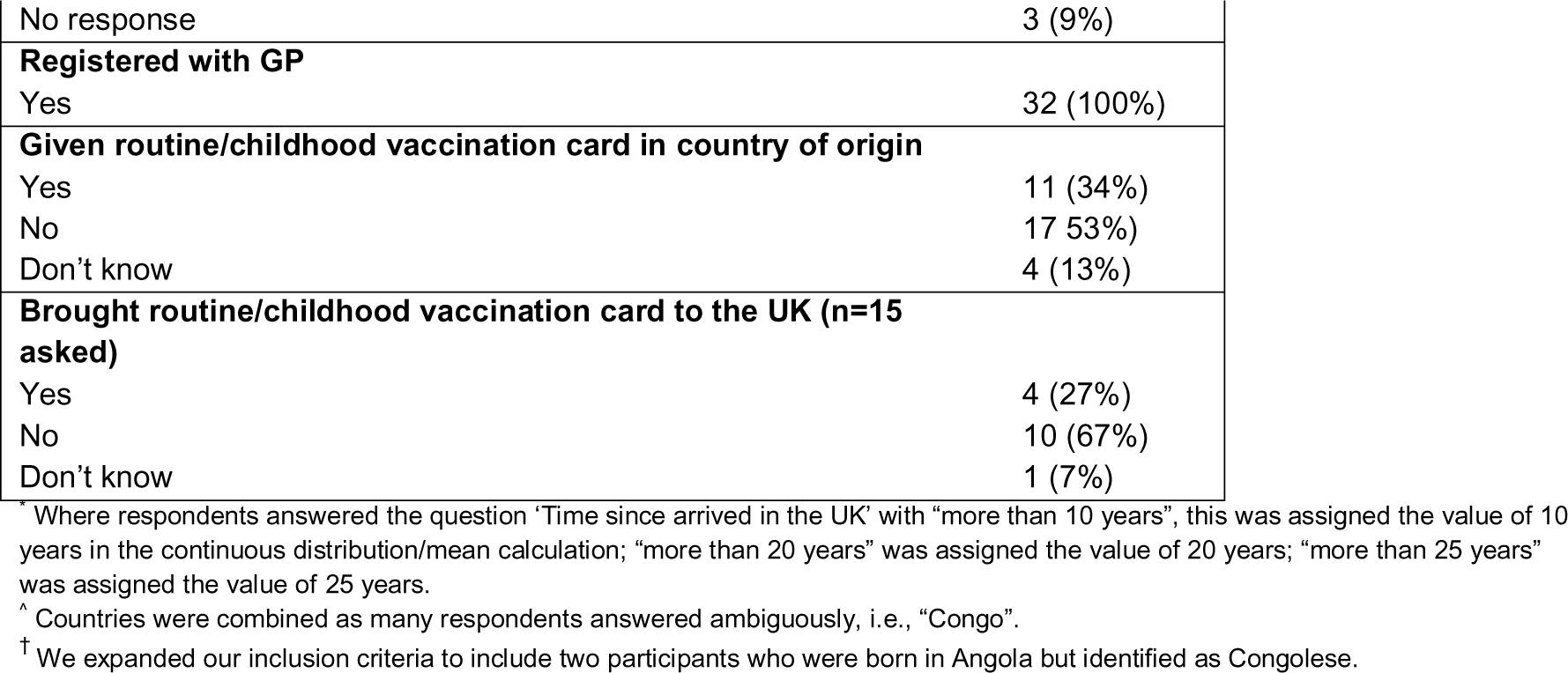
Characteristics of qualitative interview participants (n=32).

### Results part 1 – barriers and facilitators to COVID-19 vaccination and health intervention preferences

Here, we briefly report two themes: barriers and facilitators to COVID-19 vaccination and health intervention preferences. We identified 14 barrier concepts, organised under 5 topic headings (vaccine safety concerns, vaccine effectiveness concerns, vaccine necessity and norms, issues relating to information and communications, and government distrust), and 10 facilitator concepts, organised under 8 topic headings (accessibility of the vaccine, opportunity to discuss with a GP or other trusted source, higher risk perception and saliency of the disease, social influences, respect for authority, trust in government, belief in medical research process, desire to protect self and others). Examples of the data are shown in supplementary table S1.

#### Barriers

Vaccine safety concerns included uncertainty about the COVID-19 vaccine development process and speed, beliefs about consequences due to personal risk factors (e.g. blood clots), a negative experience (e.g. side effects from an earlier dose), knowledge of vaccine scares and historical events (e.g. contracting vaccine-derived poliomyelitis), or belief in rumours and conspiracy theories about the vaccine’s effects.

> *“My issue was on the blood clot side because when I had my kid, I was bleeding a lot, I lost 1 litre plus. So, when I heard on the news that people were having blood clots I said, my God, it makes me feel really scared.” – P5, female*

> *“Yes, some children have become disabled after receiving polio vaccine. […] [They are afraid] because the side effects of vaccine have caused to their children to become disabled, and they don’t want again to take the risk.” –P2, female*

There were also concerns around the vaccine’s effectiveness, and the need for multiple doses or boosters. Participants questioned the necessity of the vaccine when it doesn’t necessarily prevent infection and contrasted the COVID-19 vaccine with other vaccines such as the influenza vaccine, which they perceived to be more effective. One participant said, “I prefer flu vaccine because that one will protect you.” (P21, female)

Issues relating to information and communications were another important barrier. Many participants highlighted how language and literacy barriers had directly influenced their vaccination decisions, for example, not having access to an interpreter, or through exposure to misinformation and rumours in their informal networks, causing fear and distress.

> *“I refused [the vaccine] the first time… Because I came recently in the country, and I was not sick. I just came and I couldn’t speak English. I refused. No, I wanted to have an interpreter to explain to me…” P28, female*

> *“It was not easy for me [to get the vaccine] because there was so many rumours and I was questioned myself if do I have to take it or not. We came in this country to seek protection.” – P4, female*

A few participants also said they felt confused and overwhelmed by the official information and public health messaging, which had been complicated and at times contradictory. For example,

> *“I was scared and reluctant about the vaccines because I was confused with the information from research**….** I was not sure because scientists were not clear in their language.” – P6, female*

Widespread exposure to misinformation and rumours also made it difficult for participants to know what to believe and enhanced distrust towards authorities and public institutions. Our data suggest that many participants felt the official public health communications used by the government and NHS were coercive, and this increased their scepticism of the response, including the vaccine. Many participants said they felt they were being “forced” or “imposed” to take the vaccine, that freedom of choice had been taken away, and this had made them question the government’s motives behind the vaccination programme. For example,

> *“I have been constantly receiving letter pushing me to receive vaccine. […] I would do it voluntarily but not by force. Now they are forcing people and I don’t know what is hidden behind this vaccine?” –P16, male*

Participants voiced concerns that they might be being exploited and used as “guinea pigs” by the NHS and government and alluded to present day racism and historical events involving the exploitation of black and African populations by white Europeans. Some also commented that they felt bombarded by instructions and rules from the government and NHS about how to behave but these instructions lacked the information to help them feel safe or understand the rationale.

#### Facilitators

Most participants knew how and where to get a COVID-19 vaccine, suggesting that access was not considered a major barrier in this context. For participants who had received a COVID-19 vaccination, having the opportunity to discuss and ask questions with a GP or another trusted source, receiving encouragement and support from their social network and family, and seeing others getting vaccinated were important facilitators. The effective forms of communication that participants used to motivate their peers were in direct contrast to the coercive tone of the official messaging that participants cited as a barrier.

> *“We keep advising them not by force, but patiently to tell them respectfully, to explain to them it’s like this, it’s important, it’s for saving life, saving our kids, saving everything in our community. Something like this.” – P26, female*

#### Health intervention preferences and suggestions

Interview participants’ preferred information formats, trusted sources and messengers, communication channels, messaging contents and suggestions for interventions are summarised in Box 1.

##### Box 1. Summary of preferred health information formats, trusted sources, messaging contents, information channels and meeting points

| **Box 1. Summary of preferred health information formats, trusted sources, messaging contents, information channels and meeting points** |  |
| --- | --- |
| Formats | Oral and visual communication preferred to written formats, face-to-face, in groups or one-to-one, using Lingala and other common languages, clear and simple language |
| Trusted sources | GPs, local healthcare workers, local community organisations, friends, family, teachers |
| Information channels and meeting points | Barber shops, African food shops and restaurants, Church, parties, football/running clubs, local community organisations, a range of Congolese, African, Francophone, and social media, WhatsApp, TV and YouTube (e.g. Tele Tshangu, Top Africa) |
| Messaging contents | • Basic information to explain risks of COVID-19 and benefits of vaccination – don't just tell us to do it without explaining why<br>• Transparent information about COVID-19 vaccines and clinical trial findings<br>• Myth-busting and countering rumours and misinformation<br>• Clear, concise, and relatable messages<br>• Information about what to expect after your vaccination and how to plan accordingly (e.g. in case you feel unwell and need time off work) |
| Suggestions | • Workshops and face-to-face meetings for the community with health professionals (in Lingala or with interpreters)<br>• Increased opportunities (formal or informal) for conversation about the vaccine and sharing information about its benefits with those who are uncertain or hesitant<br>• Encouragement and motivational messaging led by and within the community<br>• Leaflets with supplementary information<br>• Sustained consultation and involvement of the community in developing future interventions |

### Results Part 2: behavioural mapping exercise and selection of interventions

Most of the barrier data related to four TDF domains: beliefs about consequences, emotion, social influences, and environmental context and resources, with smaller clusters of data related to optimism, decision-making processes, and deficits in knowledge (Table S1).

From the mapping and behavioural diagnosis exercise (31), we identified that psychological capability (specifically: knowledge; decision processes), reflective motivation (intentions; beliefs about consequences; optimism), automatic motivation (emotions/fear) and social opportunity (social influences) needed to be addressed through the intervention design. These were developed into four ‘How Might We…’ questions, for example, *How might we increase people’s knowledge about the benefits of COVID-19 vaccination and influence their decision processes?* Our chosen 5 (out of a possible 9) intervention functions for intervention development were Education, Persuasion, Modelling, Enablement, and Environmental Restructuring (the relationships between these are shown in Table S2).

A summary of possible intervention components (behaviour change techniques and mode of delivery) that were generated by the coalition in response to these questions and their corresponding intervention functions are shown in Table 3.

**Table 3.**
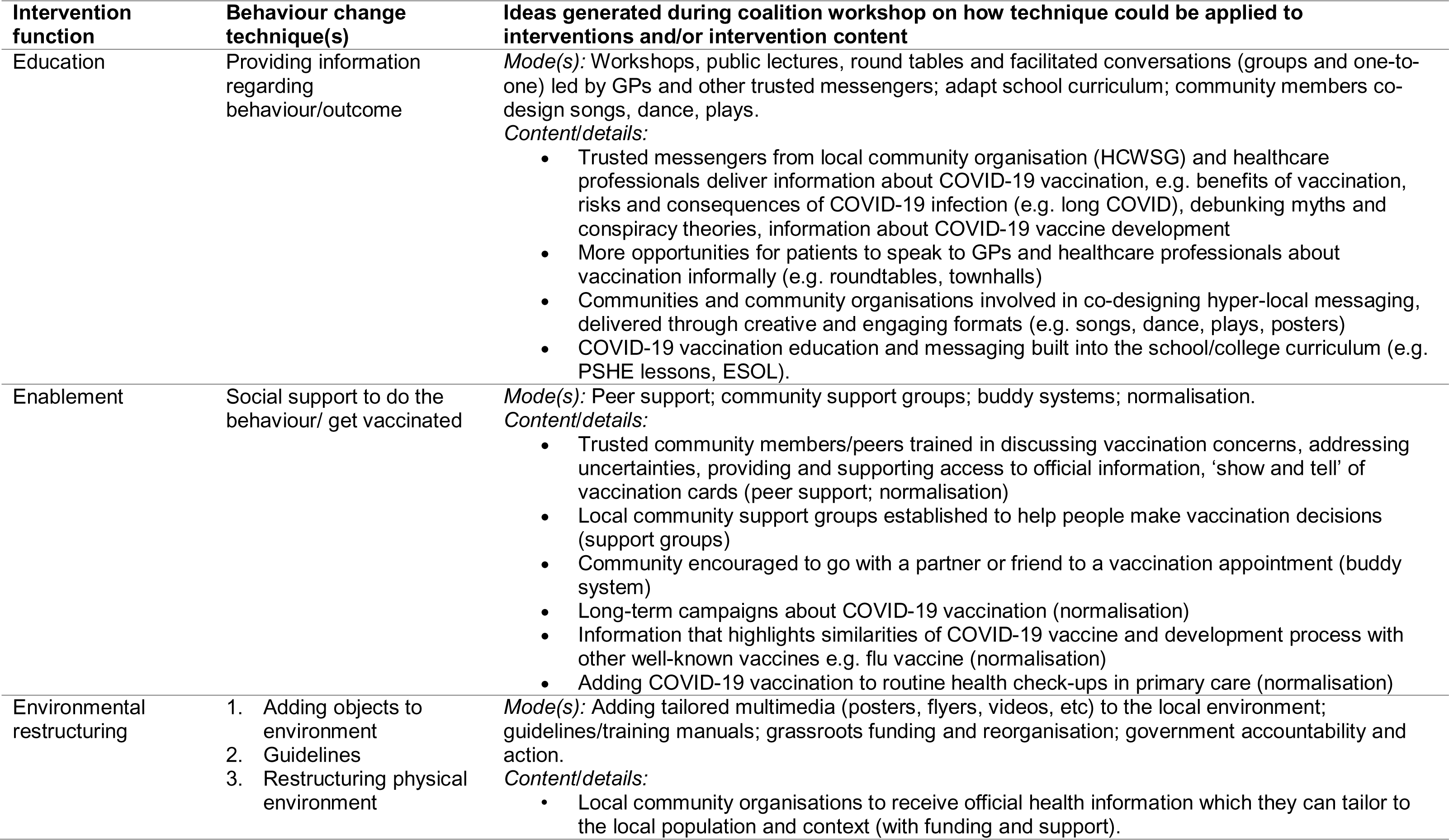
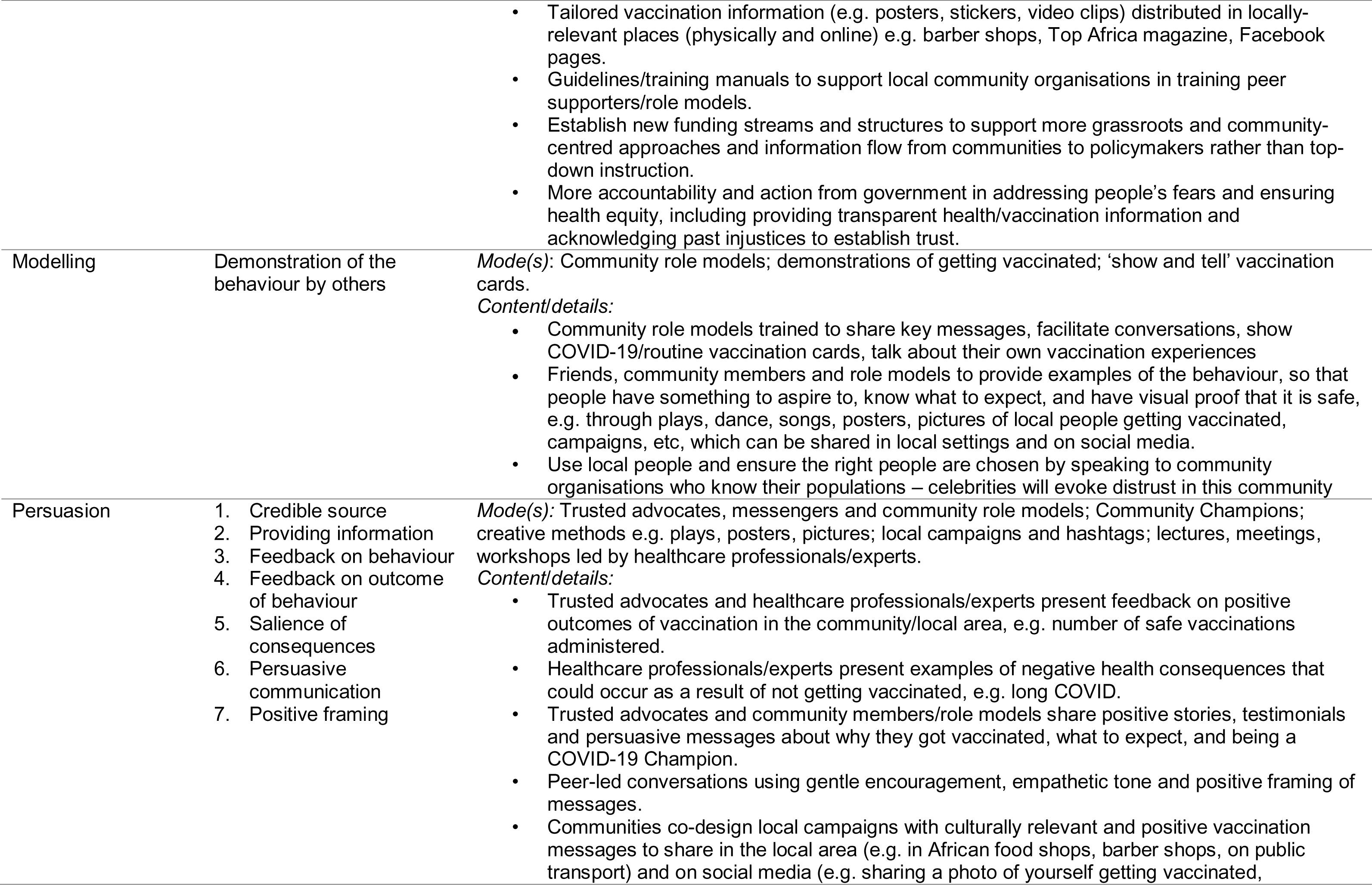
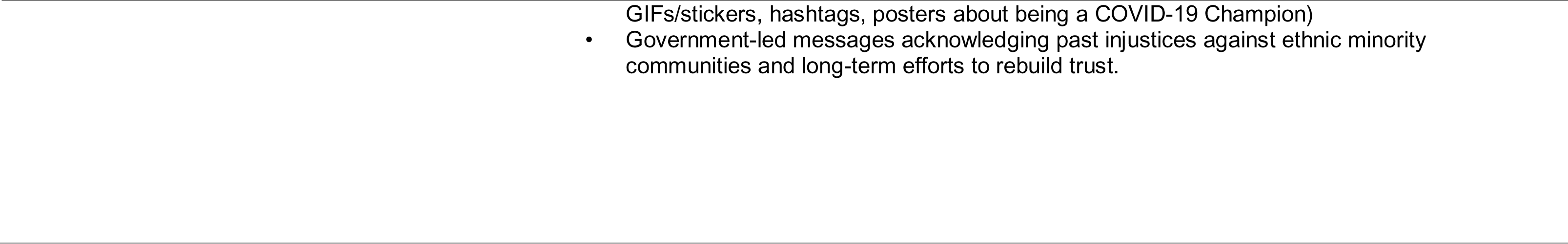
Intervention functions and potential behaviour change techniques, modes of delivery and types of content ideated during coalition workshop.

Following collective appraisal of the data, participants’ suggestions, and ideas generated in stage 4, the coalition decided by consensus on 3 intervention components to take forward to the co-design workshops:

1. Community-led workshops/meetings about COVID-19 vaccination (to increase knowledge and access to credible information, counter rumours and misinformation, address emotional and fear responses)
2. Creative storytelling/performances about COVID-19 vaccination, e.g. plays, songs, dance (to increase knowledge, model behaviour, encourage and normalise vaccination)
3. Visual media, e.g. posters, GIFs (to increase knowledge and access to credible information, counter rumours and misinformation, address emotional and fear responses)

### Results Part 3: outputs of co-design workshops

#### Intervention component 1: community-led workshops/meetings

Both groups of participants co-designed a community-led workshop/meeting plan, summarised in Table 4. The suggestions from both groups were broadly similar and complementary. Participants suggested the content should include COVID-19 information and wider health topics, including information for newly arrived migrants, and could be delivered as a series, with opportunities to ask questions, discuss and share experiences. Invited speakers should be specialists and health professionals directly involved with healthcare and vaccine development and did not need to be local or Congolese, but the sessions should be done in partnership with the local community organisation ([name redacted]). Key tailoring needs included holding meetings locally and in person where possible, using oral communication in Lingala at a minimum and with additional interpreting services if possible, and scheduled in advance, preferably on Friday or Saturday and not Sunday. Sessions should take place regularly rather than as pop-ups, as frequency and dependability are important.

**Table 4.**
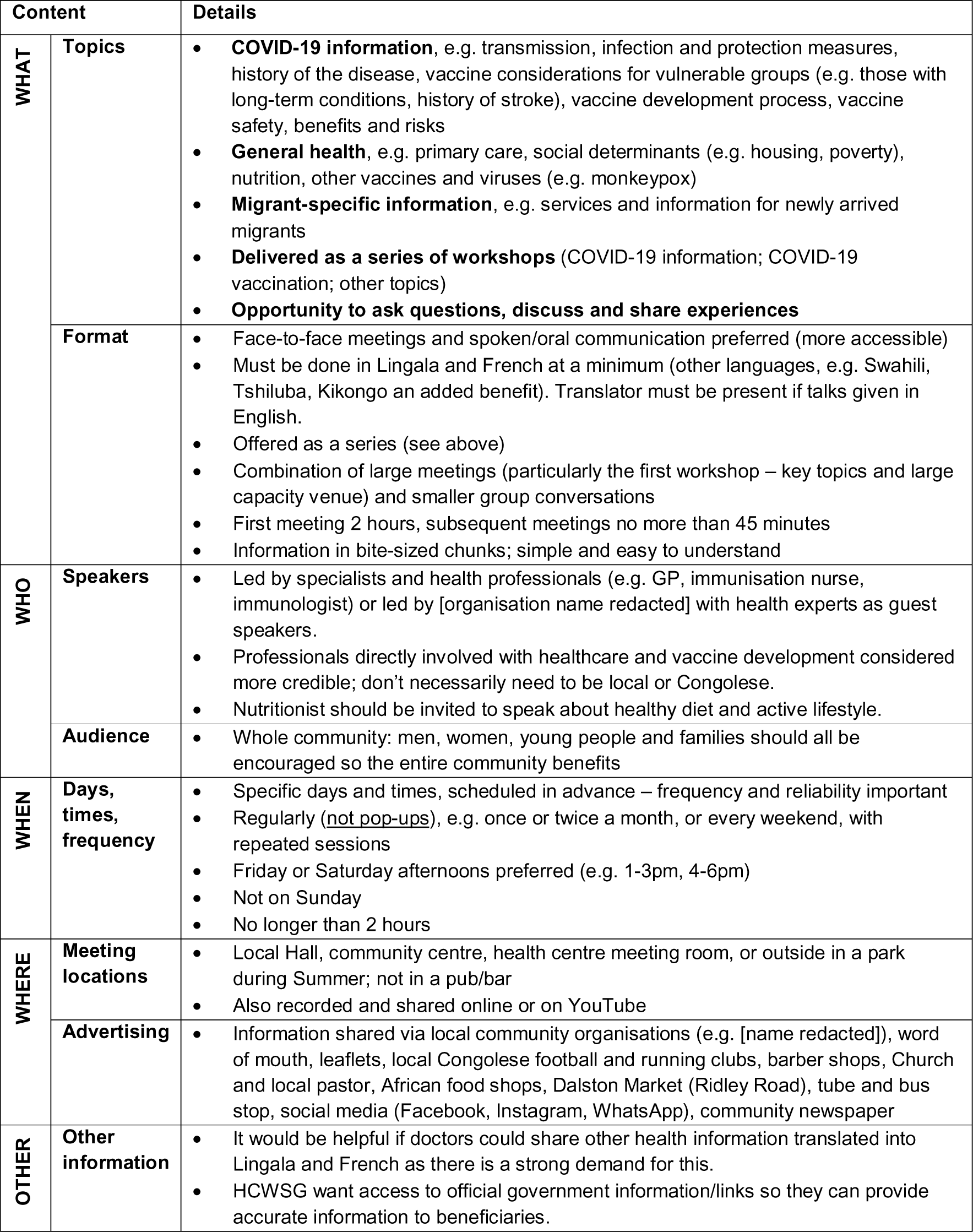
Summary of co-designed workshop/meeting content (Intervention component 1).

#### Intervention component 2: short plays

Participants chose to write short plays to address fears/misinformation and encourage their community to get vaccinated, which they sketched out using storyboards. The plays and their key messages and mechanisms of delivery are shown in Figure 2.

**Figure 2.**
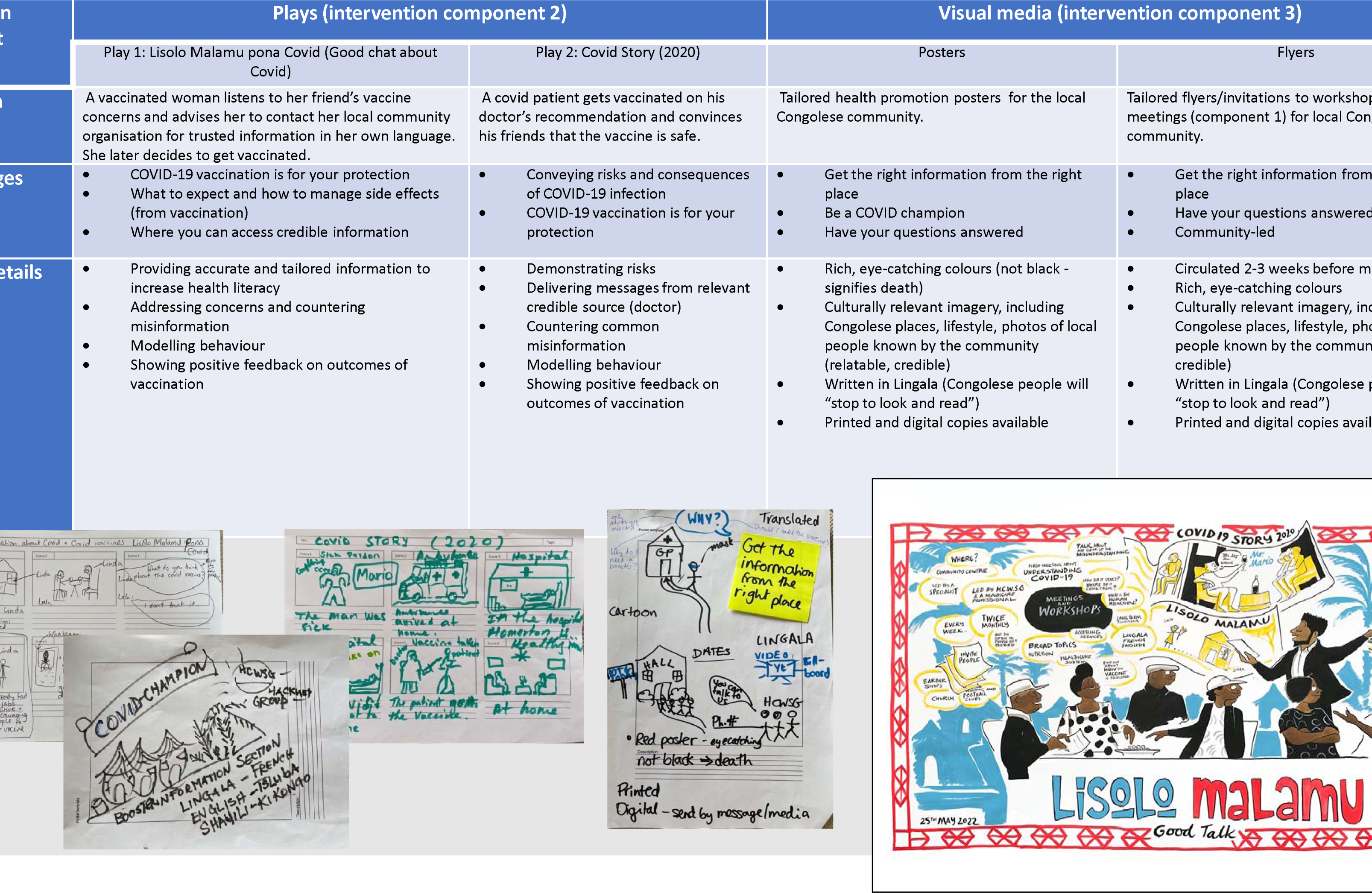
Table summarises the co-designed plays and visual media (posters and flyers) for intervention components 2 and 3, including descriptions, key messages and their delivery. The lower panel shows photographs of a selection of prototypes (storyboards and sketches) co-designed during the workshops (left) and the artist’s live drawing of the workshops and final intervention comprising workshops, plays and posters (right).

#### Intervention component 3: posters and flyers

Participants chose to make campaign-like posters about COVID-19 vaccination and flyers to invite people to the workshops/meetings co-designed in the previous activity. Examples of the posters and flyers, including key messages, content, and specific details, are shown in Figure 2.

#### Artist’s impression of workshops and intervention components (visual minutes)

The image shown in Figure 2 was drawn live by a professional artist during the co-design workshops and depicts the participatory process and the three final intervention components. This artwork has been reproduced to support funding applications and disseminate the results to the target population, key stakeholders, and local commissioners.

#### Evaluation

We received 38 completed evaluation forms from the interviews and co-design workshops. The feedback was positive and included: felt welcomed and an important part of the Congolese community, felt able to share opinions and experiences truthfully, found the discussion useful and important, and appreciated that Congolese language was recognised. One participant commented positively on the day but expressed frustration with the national vaccination guidance. Feedback on the workshops included that they were fun, sociable, well organised, and professional.

#### Dissemination

Study findings and interventions were shared with the local community and target population through a community celebration and research dissemination event in July 2022, attended by approximately 45 community members, a local councillor, and a popular YouTube news channel for African diaspora (which livestreamed the presentation of key findings to its 18,000 subscribers), and to academics and policymakers at 2 international conferences and meetings. A project results and impact brief will be shared with local and national stakeholders.

## DISCUSSION

Much COVID-19 research has focused on ethnically and racially minoritised communities with less attention given specifically to migrants. Where studies have explored migrants’ experiences of COVID-19 vaccination, few have taken a participatory line of inquiry or attempted to use those insights to co-design a vaccination intervention. Vaccination studies also less often focus on adult age groups. Our study makes a unique contribution by being the first of its kind to use a participatory and theory-informed approach to co-design a vaccination intervention with adult migrants in the UK (and an African migrant community experiencing specific barriers to accessing health interventions), with implications and lessons learned for delivering routine and catch-up vaccinations to adult migrants and co-designing other types of health interventions with marginalised migrant communities. We gathered novel insights into the beliefs and experiences related to COVID-19 vaccination of Congolese migrants in the UK and used behavioural theory, design thinking and CBPR to translate these insights into a practical, tailored, multi-component vaccination intervention for adults that was well-received by participants. Our population were primarily older adult refugees and asylum seekers, who had lived in the UK for more than 10 years and had limited English proficiency. In stage 1, we identified several key barriers to COVID-19 vaccination in this population, including concerns about COVID-19 vaccine safety, effectiveness and consequences, difficulty understanding or accessing public health information and exposure to misinformation, and scepticism stemming from pre-existing government mistrust rooted in experiences and histories of racism and discrimination. By applying an evidence-informed, integrated theoretical framework to the data, we showed that most of the barriers related to the theoretical domains of beliefs about consequences, emotion, social influences and environmental context and resources, and that an intervention that used behaviour change techniques based on education, persuasion, modelling, enablement and/or environmental restructuring (intervention functions) may be effective at achieving the target behaviour (“getting a COVID-19 vaccination”) in this population. We also identified participants’ preferred formats and sources of information, including a strong preference for oral communication using Lingala language, visual formats, and messages delivered by health professionals and local community organisations that they knew and trusted. Members of the target population subsequently co-designed three culturally relevant and tailored intervention components: community-led workshops/meetings, short plays, and posters, which aimed to increase knowledge and access to credible information, counter rumours and misinformation, address emotions and fear responses, model behaviour, and encourage and normalise COVID-19 vaccination. Conducting the study through a community-academic partnership, which meant that the study was also designed and led by members of the target population, further ensured that the findings and outputs were contextualised, appropriate and embedded within our target community.

### Intervention development

The in-depth interviews and co-design workshops revealed several important considerations for the development of interventions with Congolese migrants in this context, and for the tailoring of interventions to migrant populations in general. Below, we discuss the key findings, including the need for culturally and linguistically tailored interventions and the importance of community connectors in delivering health information and supporting local implementation and adaptation of interventions, particularly given the distrust we identified towards government-led campaigns and policies. We also discuss in detail the ways in which we tailored our interventions according to the target population’s specific values, preferences, cultures, and personal histories, and used behavioural insights to tailor our public health messages.

The preference and need for oral communication and Lingala language is an important finding and highlights the unmet linguistic needs of this population, who likely experience social exclusion and pay a “linguistic penalty” in society as a result (45). Lack of translated information is a well-documented barrier to vaccine uptake among migrants (10), while preferences for oral information have also been reported by Moroccan, Turkish and Somali migrant populations (46, 47), highlighting the need for health communication approaches that better recognise migrant populations’ linguistic and cultural diversity. We were unable to find any COVID-19 health resources in England available in Lingala, although there were some provided by Public Health Scotland, the devolved health system for Scotland (48). Translation of public health information (for example, by public institutions such as UKHSA (49)) is typically based on dominant languages, meaning larger, more established migrant populations (such as Polish, Punjabi and Bengali speakers in the UK) may be better provided for.

While pragmatic, this approach overlooks the potentially greater need for accessible resources of smaller, more marginalised populations, such as the Congolese migrants in our study. We suggest these needs and factors should be better considered by policymakers when deciding where to direct funds for linguistic and cultural adaptation of public health information, to reduce withstanding health inequities.

Participants in our study highlighted the importance of involving their trusted local community organisation in delivering the workshop intervention, in partnership with subject matter experts for credibility. These findings highlight the crucial role of local community connectors and trusted messengers in facilitating vaccination opportunities for this population. Community-based interventions and community bridging strategies involving community members as volunteers or connectors have been shown to improve connections between services and socially excluded or disadvantaged individuals and be relevant to reducing health inequalities (50). Although typically associated with low- and middle-income countries, high income countries are increasingly recognising their potential, particularly for engaging with minoritised populations (50–52). Working in partnership with people and communities is now considered “critical” by NHS England, and the recent Health and Care Act 2022 (53) was developed to enable a more collaborative system that addresses the health inequalities highlighted by the pandemic and offer more tailored and personalised care to diverse communities. A positive outcome of the pandemic is that many local authorities in the UK are now implementing ‘Community Champions’ schemes, including within ethnic minority communities disproportionately affected by the pandemic. However, it will be important to ensure that smaller organisations who are already informally supporting their communities (such as [name redacted]) are also recognised and supported through local funding and capacity building mechanisms, and that the political will and frameworks supporting these efforts are sustained long term and with ongoing community input. Local authorities will also need to be sensitive to how their actions to address inequity by focusing on larger or more dominant groups may be perceived and heighten the sense of exclusion felt by other underserved groups. Our interview participants expressed frustration that there had “never been a workshop” for their community. They also requested regular provision through a recurring series of workshops, suggesting there is both demand for and a current deficit of such information.

Our second and third intervention components used creative ways to communicate tailored health messages that were informed by the behavioural diagnosis and participants’ lived experience. Creative approaches, such as community theatre and visual art, that support preferences for oral forms of health promotion, have been successfully used in community development for decades and are well recognised tools. The participatory nature of our study, which enabled community members to participate fully in the intervention development process, meant that the resulting interventions were nuanced, culturally appropriate and specifically tailored to the local context and resonated better with the target population. The choice of plays and visual media involving representative role models as the protagonists were selected based on the target population’s preference for oral communication and popular forms of entertainment, as well as the cultural importance of storytelling. A key message participants wanted to convey through the plays was that ‘the COVID-19 vaccine is for your protection’, in response to the ubiquitous misinformation and rumours circulating and concerns about the possible negative consequences of receiving a relatively new vaccine. They did this by directly and empathetically addressing genuine concerns of their community members. For example, in the play ‘Lisolo Malamu pona Covid’, the two protagonists Lala and Linda discuss the misinformation and the emotions it evoked, before supporting each other to seek out more credible sources of information. The ways in which these messages were delivered demonstrates how behaviour change techniques, identified through a theory-informed approach, can be adapted and tailored to cultural values, beliefs, and local context, combining behavioural theory with community-led co-design processes. The plays incorporated local reference points, such as shopping at Dalston market, which is a central part of the community, and customs such as bringing a gift when visiting a friend in their home. Humour and references to day-to-day interactions introduced light relief to an otherwise serious topic, helping to make the scenarios more relatable and personal, which can help promote a sense of representation and belonging (54). For example, we saw Lala exclaiming “Now I can go to Nando’s!” (a popular chicken restaurant) after receiving her COVID-19 vaccination pass, and Mr Mario enjoying a glass of wine at home after experiencing no adverse events following his COVID-19 vaccine. These references also aligned with important moral values of the target population, such as agency and liberty (see upcoming manuscript, *in preparation*), which evidence suggests may promote better engagement with the subject matter and uptake of the target behaviour (55).

The key messages of the posters were ‘get the right information from the right place’, aiming to signpost people to credible information, answer common questions, and debunk rumours and misinformation, and ‘be a COVID-19 champion’, reflecting the role individuals within the community can play in supporting and promoting vaccination uptake. The first message speaks to the vast misinformation participants reported and their confusion and anxiety trying to navigate information and make vaccination decisions. The latter message links closely to the communal nature of the population’s society and the social influences identified as facilitators in the qualitative study (particularly positive outcomes feedback, seeing others getting vaccinated, and positive reinforcement via community members) and participating in the vaccine programme to protect oneself, family, and the wider community. Contrary to the numerous celebrity-fronted public information campaigns during the COVID-19 pandemic (56, 57), participants made clear they did not want to see images of celebrities used to promote vaccination, as this would evoke distrust, but preferred to see people from their communities participating in the vaccine programme, as this would be more trustworthy and relatable. Culturally relevant imagery and rich colours were also considered important and a strong feature of each of the designs. Adding these details may be a simple way to make a campaign stand out to a specific target population and feel more inclusive, particularly for underserved and minoritised migrant groups.

### Evaluation and impact of participatory approach

A goal of participatory research is to reinforce local capacity and solutions and promote transformative change (34, 58, 59). Through this study we enhanced community capacity through our approach to sharing power, recognising and celebrating community assets and expertise, and by providing skills-based training and leadership opportunities for community partners. This focus on community assets and the ability of communities to find their own solutions is contrary to deficit models which suggest that barriers are due to difficulties around language and other issues relating to access and trust.

Using participatory methods, we demonstrate that underserved communities, such as migrants, are resilient, resourceful, and use community assets to find real-world solutions to their health needs. Planning the study through an equity lens facilitated participation and resulted in high engagement and strengthened the community organisation’s [name redacted] links with more marginalised members of their community. Our partnership with the umbrella organisation [name redacted] and relevant expertise in our coalition [initials redacted] meant that we also ensured participants who attended the study activities were signposted to relevant services, such as healthcare, housing, and education courses. We accelerated translation of research to action by co-designing relevant interventions driven by local values and needs. Without our participatory approach, which enabled community members to participate with full voice, many of our findings may not have been uncovered nor translated into viable, specific solutions. The interventions and artwork created are meaningful to the local community organisation and participants, as they represent and value their participation, lived experience and the knowledge that was co-created through their involvement in the study. The artwork has been of clear benefit to the coalition and specifically the local community organisation (HCWSG), who have used it as a visual aid in meetings with stakeholders and to support funding applications. Anecdotal evidence also suggests that providing a friendly, non-judgmental space to talk about COVID-19 vaccination with trusted members of the community through our community days increased uptake of the vaccination in many people who were initially hesitant, demonstrating the power of a community-engaged, participatory approach.

### Limitations and strengths

Naturally, our study has some limitations. Because the intervention design was informed by behavioural theory, the interventions focus on modifiable behaviours and do not directly tackle structural barriers such as systemic racism and discrimination which are thought to play a fundamental role in ethnic inequities in vaccine hesitancy (60). However, through our in-depth qualitative analysis and engagement we successfully explored the social, societal, and contextual factors relevant for this population and ensured they were reflected in the tailored content, and using trusted voices ensured that the interventions were considered relevant and appropriate. We also address these factors in greater depth through the reflexive thematic analysis of our qualitative data and formulation of policy recommendations at multiple socio-ecological levels in our upcoming paper (manuscript *in preparation*). Although we have not yet formally implemented and tested our interventions, our community partner has received further fundraising and capacity building support locally to enable them to continue building on this work and beyond. Future studies could mitigate high translation costs by adapting/condensing the study design to focus solely on intervention development and using a rapid data collection approach to identify intervention functions (omitting wider contextual data), however our more thorough approach arguably led to richer data that may be used to aid implementation. Translation needs could be overcome entirely by employing a study team proficient in the language of participants, though this may not always be realistic. The DRC is home to over 200 ethnic groups; however, we did not collect data on ethnic group of our participants, and at times lacked clarity on participants’ place of birth (Republic of Congo or DRC). This is a limitation of our study which we would strive to rectify in future research involving diverse diasporic communities. As such, we should be mindful that our results may reflect the views of potentially diverse ethnic groups with distinct beliefs, lived experiences and practices within a broad overarching group of Congolese diaspora, rather than a single community. A key strength of our study was its commitment to co-production and reflexivity. Inadequate inclusion of migrants in developing health interventions has been identified as a shortcoming of existing participatory health research with migrants (35). By contrast, our approach valued lived experience, embraced community members as equal partners, and empowered them to participate with full voice throughout the study (with all aspects designed and implemented collaboratively). Our generous timeline and budget facilitated building trust and community capacity and allowed for flexibility in our approach, which is critical for co-design and community-based research. We began engaging with the local community and building relationships with local partners prior to the study’s inception, which led to a trusting and productive working relationship. By involving individuals with lived experience and diverse skills and expertise in our coalition, we designed and implemented a relevant, rigorous, and well-planned study. We provided details of our budget for transparency and to highlight unforeseen costs for others wishing to conduct similar research. Although a participatory approach is labour intensive, we argue that it can lead to better outputs and outcomes than traditional approaches. Participants commented in the evaluations that they felt included, visible, and appreciated, and enjoyed the participatory process. This led to the co-design of nuanced and tailored interventions, as well as wider transformation, including personal development of community members and an increased sense of inclusion and appreciation in an underserved and minoritised community. Realistically, we must admit that power imbalances still existed in our approach, however we strived for full participation at all stages and no doubt achieved this more successfully than a study built on a solely traditional research paradigm. Finally, our study funded a black-led organisation to lead community-based research addressing issues important to their community and provided personal development opportunities to build community capacity. An alarmingly low number of black-led organisations were awarded funding in the COVID-19 response and in the community and voluntary sector in general (61–65), therefore our study perhaps makes an important contribution towards showing how community engagement and participatory research can be used to advance equity in migrant and ethnic health and health research, and dismantling the power structures creating barriers to vaccine uptake and perpetuating harm to these communities.

## CONCLUSIONS AND NEXT STEPS

The worse health outcomes of adult migrant populations during the COVID-19 pandemic and their widely reported barriers to COVID-19 vaccination have warranted the exploration into more tailored interventions to increase vaccine uptake, which consider local context, including personal histories, power dynamics, preferences and needs, and are developed and implemented in close collaboration with the target population. They have also highlighted wider inequalities and prompted research into ways of better engaging underserved adult groups specifically in vaccination campaigns, learnings from which can be adapted and used for strengthening routine immunisation programmes. This study reports on the theory-informed co-design of a tailored COVID-19 vaccination intervention to address these complex challenges in an underserved Congolese migrant population in London. Our findings and co-designed intervention demonstrate how alternative formats and delivery mechanisms, including linguistic adaptations and use of trusted community connectors, can be used to successfully disseminate health information to intersectionally marginalised populations, and how behavioural theory combined with local and cultural insights can help to develop tailored messages which resonate with migrant communities. These adaptations and designs were simple and feasible to produce, acceptable to the target population, and are likely to be relatively low-cost to implement. Importantly, participants were positive about their involvement and said in the evaluations they felt the study was inclusive and made them feel valued. The participatory approach of the study (reported elsewhere (36)) offers a replicable model for engaging with underserved communities in an empowering and equitable way, demonstrating how academic and community partners can better foster mutual exchange of expertise and work effectively together outside of traditional power structures. Next steps will involve refining, implementing, and testing the interventions, and potentially adapting and expanding the content to routine vaccinations and wider health needs, as requested by study participants and to address gaps exacerbated by the pandemic. The findings also hold relevance to the co-development and implementation of other health interventions and health promotion activities with these and other similar communities. Future research should build on this empowering approach to engaging with underserved migrant communities, with the goal of developing more sensitive vaccination services and interventions which respond to migrant communities’ unique needs and realities.

## Declarations

### Ethics approval and consent to participate

Ethics was granted by the XXX Research Ethics Committee (REC reference 2021.0128). All participants provided informed consent and were older than 18 years at the time of recruitment to the study.

### Consent for publication

Not applicable.

### Availability of data and materials

Data will be available from the corresponding author on reasonable request.

## Authors’ contributions

[initials redacted] had the initial idea for this research study. [initials redacted] jointly conceptualised and conducted the study, including investigation, recruitment, analysis, and project administration. [initials redacted] trained the study team as peer researchers, managed the study and budget, and wrote the original draft. [initials redacted] provided supervisory support and advice during the study. All authors [initials redacted] supported the development and refinement of study tools and procedures. All authors [initials redacted] read and approved the final manuscript.

## Data Availability

All data produced in the present study are available upon reasonable request to the authors.

## Acknowledgments

We are grateful to the participants who shared their life experiences with us and actively participated in the study. We are also grateful to our coalition for their dedication and commitment to sharing knowledge, working and learning together for the betterment of community health. We thank the local community organisations, including [names redacted] and artist [name redacted], that championed our project and generously provided spaces and resources to support our work. We also thank our [name redacted] Project Board, particularly [name redacted], for their advice and support.

