## Supplemental Table 1, Supplemental Table 2 for "Co-designing a theory-informed, multi-component intervention to increase vaccine uptake with Congolese migrants: a qualitative, community-based participatory research study"

**Supplementary tables**

Table S1. Examples of barriers and facilitators to COVID-19 vaccination identified through in-depth interviews with Congolese migrants, mapped to TDF domains.

| BARRIERS |  |  |  |
| --- | --- | --- | --- |
| Examples of data | Topic heading | Description of key barriers | TDF domains |
| “For the COVID vaccine, so many people in the community are reluctant to have it because they are not sure if by the time it was produced to experimental level was it secure enough for people to have it.” – P5 (Female, 50-64 years) | Vaccine safety concerns | Uncertainty about the vaccine development process and speed | Beliefs about consequences |
| “My issue was on the blood clot side because when I had my kid, I was bleeding a lot, I lost 1 litre plus. So, when I heard on the news that people were having blood clots I said, my God, it makes me feel really scared.” – P5 (Female, 50-64 years) | Vaccine safety concerns | Concerned about consequences due to personal risk factors (e.g. blood clots, pregnancy) | Beliefs about consequences; Emotion |
| “Because of the reaction that I had [to the COVID-19 vaccine] I will never take another because I was so scared that I am going to die.” – P32 (Female, 25-49 years) | Vaccine safety concerns | Concerned about consequences due to adverse event/side effects of first dose | Beliefs about consequences; Emotion |
| “There were rumours saying that those vaccines will reduce 10 years from your age because they want to reduce the number of people in the world. This vaccination is very bad.” – P18 (Male, 25-49 years)  “They are listening from YouTube that is the reason people are scared… They are saying if you have it you my die, if you are a young girl you may not have children and if you old you will not live longer.” P27 (Female, 65+) | Vaccine safety concerns | Exposure to rumours and conspiracy theories; belief in misinformation | Beliefs about consequences; Emotion; Knowledge; Social Influences |
| “Yes, some children have become disabled after receiving polio vaccine. […] [They are fearful now] because the side effects of vaccine have caused to their children to become disabled, and they don’t want again to take the risk.” –P2 (Female, 25-49 years)  “Can I give a testimony about a child back in DRC. I have seen when the girl was born and grew up to the age of 10-13 years old who received polio vaccine; surprisingly, the girl has become disable due to the side effects of polio vaccine. […] I am a living witness about what happen to this child after receiving polio vaccine.” P16 (Male, 65+ years) | Vaccine safety concerns | Apprehension following vaccine scares/historical events (e.g. vaccine derived polio and paralysis) | Beliefs about consequences; Emotion |
| “How can you explain that people are dying even they have been vaccinated?” P4 (Female, 50-64 years)  “The vaccine prevents you not to catch illness, but it doesn’t means that you may not be infected. There were a lot of confusion, and this was the reason I was not ready to be vaccinated.” – P6 (Female, 25-49 years)  “Yes, [I have] flu vaccine, every year. I prefer flu vaccine because that one will protect you.” P21 (Female, 50-64 years) | Vaccine effectiveness concerns | Not confident that the COVID-19 vaccine is effective or protective | Beliefs about consequences; Knowledge; Optimism |
| “But, COVID-19 vaccines have created many problems, and you need to take 3 to 4 times.” – P20 (Female, 65+ years)  “I was wondering about all these vaccines. The GP said I must take the booster jab as well.” – P14 (Male, 50-64 years) | Vaccine necessity and norms | Perception of risk and necessity of multiple doses | Beliefs about consequences; Emotion; Knowledge |
| “No, vaccine is something that you take once a year but now it has become too much and this is causing a lot of illnesses.” – P20 (Female, 65+ years) | Vaccine necessity and norms | Number of required doses does not align with understanding of vaccination/norms | Social influences; Decision processes; Knowledge |
| “Our antibody are strong comparing to people of this country they should receive vaccine first than us. For myself, I have been living in UK for many years and I never received any vaccine but I am feeling fine, and healthy.” -P16 (Male, 65+ years) | Vaccine necessity and norms | Preference for natural immunity; perceptions of immunity spectrum | Knowledge; Social influences; Decision processes |
| “People are confused about the spread of the wrong information from doctors and media. There is a lot of confusion with different information. Doctors are contradicting with their information. We don’t know if we have to take it or not. “ – P4 (Female, 50-64 years)  “I was scared and reluctant about the vaccines because I was confused with the information from research…. I was not sure because scientists were not clear in their language.” – P6 (Female, 25-49) | Issues relating to information and communications | Confused by public health messaging; inconsistent messaging | Knowledge; Environmental context and resources |
| “It was not easy for me [to get the vaccine] because there was so many rumours and I was questioned myself if do I have to take it or not. We came in this country to seek protection.” – P4 (Female, 50-64 years) | Issues relating to information and communications | Exposure to misinformation and rumours | Beliefs about consequences; Emotion; Knowledge; Social Influences |
| “I refused [the vaccine] the first time… Because I came recently in the country, and I was not sick. I just came and I couldn’t speak English. I refused. No, I wanted to have an interpreter to explain to me…” P28 (Female, 50-64 years)  “You can give me a leaflet, and if I don’t know the language what is the point?” – P5 (Female, 50-64 years)  “There’s never been a workshop convincing people why they should take the vaccine. Especially in my community, we don’t speak English and people have got difficulty understanding information even when it’s going through TV.” - P5 (Female, 50-64 years) | Issues relating to information and communications | Unable to access information; information and language barriers | Knowledge; Environmental context and resources |
| “Also, I am questioning myself what is hidden behind COVID-19 vaccine? You know why people are refusing to be vaccinated because they are forcing people to be vaccinated like there is something behind this vaccination programme.” – P4 (Female, 50-64 years) | Government distrust | Distrust in UK government/ government’s motives; value freedom of choice | Beliefs about consequences; Emotion; Optimism; Environmental context and resources |
| “In Africa, it doesn’t matter for them. [The government] doesn’t work for their population. It stop. It works for themself. […] That’s why people in Africa, they didn’t trust [the vaccine].” – P29 (Male, 50-64 years) | Government distrust | Distrust in government of home country/region | Beliefs about consequences; Social influences; Optimism; Environmental context and resources |
| “And the third [thing I’ve heard] is that they are using us as guinea pigs and that’s a big problem.” P5 (Female, 50-64 years)  “They were rumours that the NHS is using their vaccines for test on us.” – P12 (Male, 25-49 years) | Government distrust | Inequality-driven distrust; aware of past injustices and don’t want to be exploited | Beliefs about consequences; Emotion; Optimism; Environmental context and resources |
| FACILITATORS |  |  |  |
| Examples of data | **Topic heading** | **Description of key facilitators** | **TDF domain** |
| “I did not do anything. You just turn up at the vaccination centre.” – P2 (Female, 25-49 years)  “I booked myself through NHS website and I went to receive my first dose.” – P6 (Female, 25-49 years)  “My GP surgery booked the appointment and they also sent the address where I have to take my vaccine.” – P14 (Male, 50-64 years) | Accessibility of the vaccine | Ease of access, including knowing how to book appointment or where to get a vaccine, and being able to access healthcare/primary care | n/a |
| “Yes, apart from TV and email messages, we did have 1 to 1 talking with my manager who explained about COVID-19 vaccine. The manager explained to us the advantages and disadvantages of receiving vaccines. She also advised to approach my GP.” –P12 (Male, 25-49 years)   “Like I said to you I don't trust anyone. The one [vaccine] I've taken, they said to me it’s fine because I spoke to my GP for a long time just like I spoke to you. The advantages and the disadvantages, after he explained to me I decided to take [vaccine].” P21 (Female, 50-64 years) | Opportunity to discuss with a GP or other trusted source | Having a dialogue with a trusted person – e.g. GP, employer, etc who could provide and help access credible information | n/a |
| “I have taken already 2 vaccines and I have an appointment for the booster. It is good for your own protection. I have lost one of my close friends [from COVID-19].” – P18 (Male, 25-49 years)  “Yes. Some people say it's like many thing inside you also will not walk properly, we damage you, something like this. I said, okay, it's your conviction. For me it's not to kill because if they can't kill all the world, they can't kill all the London people. It's to save us from the infection of COVID. I'm deciding, I've got my appointment, I will get it. If you people say to die, better to die with my injection COVID, than to die with COVID.” – P26 (Female, 25-49 years)  “I just went to receive my vaccine because the news was spread all over the world how people were dying.” P14 (Male, 50-64 years) | Higher risk perception and saliency of the disease | Perceiving COVID-19 risk being high and vaccines to offer protection | n/a |
| “But now covid-19 is in control and there are vaccinations, people are campaigning to encourage people to be vaccinated.” – P18 (Male, 25-49 years)  “This year, I’m telling you, we can say well done for the vaccine. You can see the people coming, they have had the vaccine and you can say I’m vaccinated for a long time and I’ll get two or three that get COVID again and they are good, less coughing and a little fever. You see the antibody working, that’s why I say for vaccination is good.” – P13 (Female, 25-49 years) | Social influences | Sense things are improving since vaccine was introduced; positive outcomes feedback | n/a |
| “After I have seen too, some people around me, they got the vaccination and they advise me. I say, okay, I need to go get [it]. […] My wife too, got the vaccination. I send her to go get the vaccination after I got my one.” P29 (Male, 50-64 years) | Social influences | Seeing others getting vaccinated; family support | n/a |
| “Some people, they’re hesitating to get the vaccination. He wants to take it but he’s scared and they say ‘I will be sick’ and they say ‘I will be down’ and all these things. And I say no, go and get it. […] I said I’ve got mine and I’m in good health, and they go there and they take it.” – P9 (Female, 50-64 years)  “We keep advising them not by force, but patiently to tell them respectfully, to explain to them it's like this, it's important, it's for saving life, saving our kids, saving everything in our community. Something like this.” – P26 (Female, 25-49 years) | Social influences | Positive reinforcement and messaging from within own community (role models, messages that promote relevant moral values e.g. liberty) |  |
| “We are here and we live here we have to follow them [the rules].” – P18 (Male, 25-49 years)  “It was an obligation to receive vaccine and I did it.” – P20 (Female, 65+ years) | Respect for authority | Tendency to follow rules/laws | n/a |
| “Some for them, it’s a scare. But I advised them, I said to them, it’s nothing. Because look at it. People is thinking wrong. They say maybe it’s a poison. I have advised them. I said, no. Look at me. In England, it’s not like Africa. Because the government for England is work for their people. Is work for their community.” P29 (Male, 50-64 years) | Trust in the government | Feeling the government has their best interests in mind | n/a |
| “For white people they will always test if it is good to give to people otherwise they can kill so many people there is not a problem for me.” P31 (Female, 25-49) | Belief in medical research process (including racial injustices) | Perception that a vaccine developed for white people will have been tested properly; higher ethical standards for white populations | n/a |
| “I received a letter from NHS inviting vulnerable people to be vaccinated and I am working with vulnerable people, I decided to be vaccinated so that I can be protected and protect other people as well.” P12 (Male, 25-49 years) | Desire to protect self and others | Getting the vaccination for protection | n/a |

Table S2. Relationship between HMW questions, COM-B and TDF domains, and intervention functions.

| ‘How might we…?’ (HMW) question | Further details and aims behind question | Corresponding COM-B domain (TDF domain) | Possible intervention functions (chosen functions in bold) |
| --- | --- | --- | --- |
| *How might we increase people’s knowledge about the benefits of COVID-19 vaccination and influence their decision processes?* | - Help them understand benefits of COVID-19 vaccination, risks of COVID-19 infection - Help them plan to attend appointments | Psychological capability (knowledge, decision processes) | **Education**, Training, **Enablement** |
| *How might we change people’s beliefs about the consequences of COVID-19 vaccination and increase their intentions to get vaccinated?* | - Help them intend to and prioritise getting vaccinated - Want to do it - Feel it is important - Believe it would be a good thing to do | Reflective motivation (intentions; beliefs about consequences; optimism) | **Education**, **Persuasion**, Incentivisation, Coercion |
| *How might we reduce people’s fears about COVID-19 vaccines/vaccination?* | - Help them to not fear COVID-19 vaccination and be able to quash rumours - Help them establish COVID-19 vaccination routines | Automatic motivation (emotions/fear) | **Persuasion**, Incentivisation, Coercion, Training, **Environmental restructuring**, **Modelling**, **Enablement** |
| *How might we create more social opportunities for COVID-19 vaccination?* | - Help them see COVID-19 vaccination as socially acceptable and expected - Others are doing it - Social support to do it | Social opportunity (social influences) | Restriction, **Environmental restructuring**, **Modelling**, **Enablement** |
